# Early development of cortical networks is modulated by Family Nurture Intervention: a multicenter replication study

**DOI:** 10.64898/2025.12.05.25341717

**Authors:** Pauliina Yrjölä, Michael M. Myers, Martha G. Welch, Sampsa Vanhatalo, Anton Tokariev

**Author notes:** **Correspondence:** Pauliina Yrjölä, Department of Physiology, Biomedicum 1, BABA center, room B129b, University of Helsinki, P.O.Box 63 (Haartmaninkatu 8), 00014 University of Helsinki, Finland.

## Abstract

Premature birth is associated with a high risk of abnormal neurodevelopment, inflicting lifelong neurocognitive consequences with major public health importance. Environmental enrichment in neonatal intensive care units (NICU) may improve outcomes, including early development of functional brain networks. Here, we studied how well the previously found brain effects of Family Nurture Intervention (FNI) are replicated in an independent trial, and when these FNI-affected effects arise in brain network development. We compared cortical activity networks in infants following FNI treatment to infants receiving standard NICU care (SC) and to term-born healthy controls (HC). We found that the prior network results, *i.e.* a decrease in the strength of phase-based networks of FNI infants that rendered them comparable to the networks of HCs, replicated well in an independent trial. Moreover, longitudinal tracking of cortical networks disclosed an early divergence of trajectories between the FNI and SC groups during the preterm period. The findings together support the idea that FNI may modulate early developmental trajectories of the cortical networks and mitigate the effects of prematurity.

## Introduction

Prematurity has been associated with changes in structural ^1–4^ and functional ^5–9^ brain network development with long-lasting neurocognitive sequelae ^10,11^. Preterm infants spend much of the third trimester *ex utero* in the neonatal intensive care unit (NICU). During this time the brain undergoes dynamic self-organization of both structural and functional networks, guided by endogenous neural activity ^12–15^ that is particularly susceptible to effects associated with medical adversities and NICU treatments ^16–18^. Many NICUs are now striving to optimize neurocritical care via minimizing care-related stress and implementing various environmental enrichment (EE) strategies ^19–21^, including human voice and music therapy ^22–24^, as well as massage therapy^25^. However, there is scarce objective and evidence-based assessment of the replicability of these strategies’ neuronal effects through early neurodevelopment, including the effects on functional brain networks needed to support later neurocognitive performance.

Comparisons across different EE strategies point to multisensory approaches as the most effective for supporting holistic neurodevelopment ^19^. A well-studied example of multisensory EE is Family Nurture Intervention (FNI) that aims to facilitate the emotional connection between mother and infant during the NICU stay ^26^. FNI leverages more than a physical contact alone (cf. skin-to-skin care or family-centered care ^27–29^) by recruiting deep emotional engagement of the mother-infant dyad. FNI has been shown to improve mother-infant coregulation ^26,30^, infant’s autonomic regulation ^31^, and physiological stability ^32^, as well as contribute to many neurobehavioral benefits ^33,34^ and changes in brain function ^35–39^. Recently, we showed that FNI also modulates the development of the infants’ cortical activity networks at term age, rendering them comparable to those of healthy term-born infants and correlating with later neurodevelopmental outcomes ^38^.

The maturation of the developing brain to achieve higher-level cognitive abilities ultimately depends on the efficient organization of cortical activity networks ^40–43^, which can be characterized using measures of neuronal interaction between brain regions, *i.e.* functional connectivity. In infants, these interactions are most readily estimated as phase-phase correlations (PPC) from recordings of electroencephalography (EEG) and are considered to reflect the coordination of peak excitability between neuronal populations on a sub-second timescale ^44,45^. Prematurity and the ensuing stay in the NICU hit the core of brain development affecting both the white matter structures needed for the cortico-cortical networks ^2,3^, as well as the early endogenous activity needed for activity-dependent organization. These effects together lead to altered neurodevelopment ^46^ reflected also on its readout, functional cortical networks ^5,6^. Here we come to a critical question: can the adverse effects on functional connectivity be mitigated by an EE intervention administered in the NICU as suggested by our recent work ^38^? Furthermore, when do these changes emerge in the development of functional networks?

In this work, we set out i) to undertake a systematic replication study of our previous findings on the effect of FNI on the functional brain networks of preterm infants, and ii) to study in more detail the developmental trajectory of these networks from preterm to term age. We hypothesized that FNI would lead to frequency-specific changes in cortical connectivity at term age, observed previously ^38^. We investigated the cortical-level PPC in high-density EEG recordings from a multisite replication randomized controlled trial, where infants were assigned to receive either FNI or standard care (SC). We examined the spatial similarity of the FNI-affected networks between prior and current study cohorts, and we compared their strengths to an external cohort of healthy controls (HC). Finally, we contrasted the developmental trajectories of functional connectivity from 33 to 45 weeks age between the two groups of age to investigate when the network development diverges.

## Methods

The main goal of this work was to assess the replicability of previously found network effects of FNI ^38^ in data from a multisite replication randomized controlled trial (RCT2) against benchmarks established in the original FNI trial (RCT1). The primary tests were statistical comparisons of group difference (FNI vs SC) in functional connectivity assessed by phasephase correlations (PPC). Next, the group difference networks’ spatio-spectral properties were compared, and network strength of the treatment groups was contrasted to an independent group of term-born HCs. Finally, the timing of the network differences was analyzed by assessing the development of these networks from preterm to term age. The overall study pipeline is presented in Fig. 1 and detailed in the following sections.

**Fig. 1.**
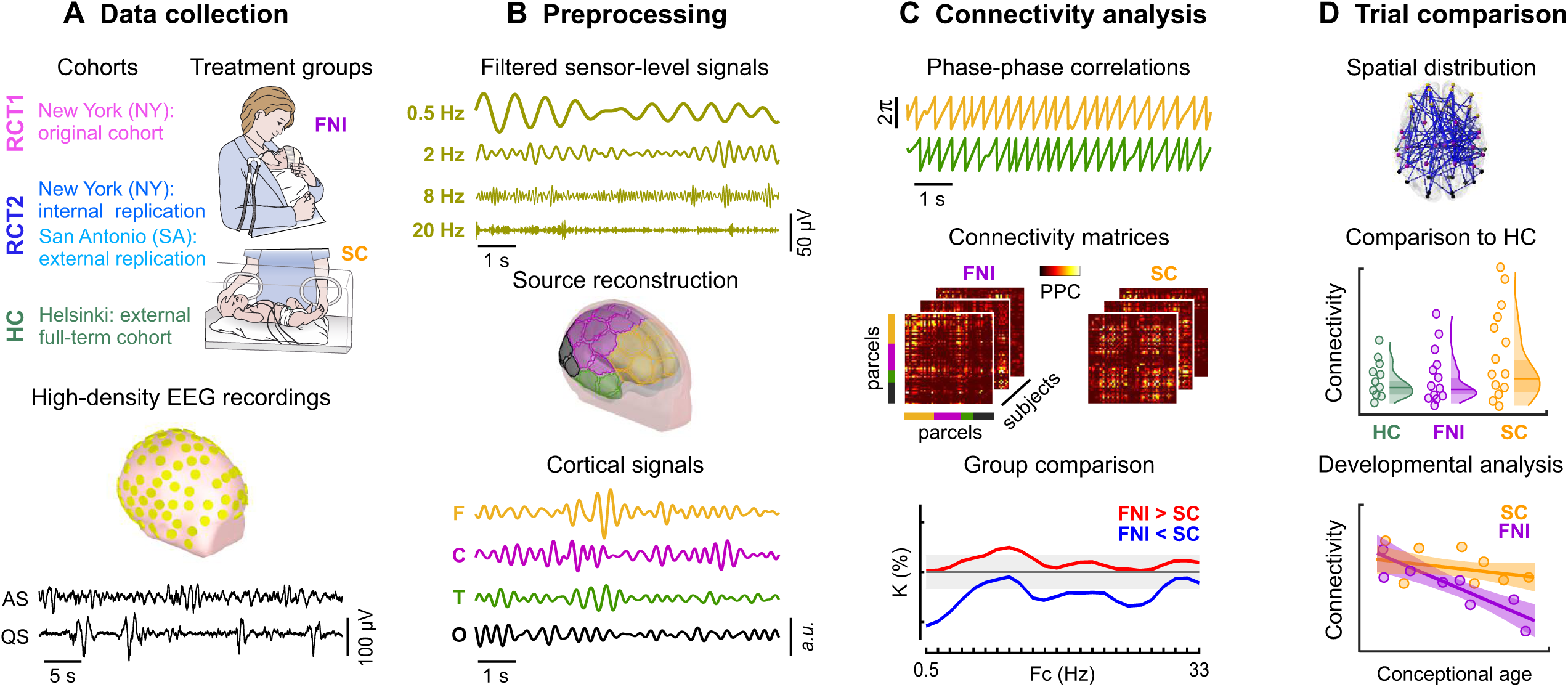
Overview of the study design. **(A)** The data used for the present study came from two randomized controlled trials of Family Nurture Intervention (FNI) of Columbia University Medical Center: RCT1 (original trial) and RCT2 (replication trial), which further consisted of two cohorts: RCT2-NY (within-site replication cohort) and RCT2-SA (external validation cohort). Preterm infants were randomly selected to receive either FNI, where mothers were facilitated in establishing an early emotional connection, or standard care (SC). An additional cohort of term-born healthy controls (HC) was obtained at Helsinki University Central Hospital. Electroencephalography (EEG) was recorded during daytime sleep with a 128-channel (RCT1, RCT2) or 19-channel (HC) cap, followed by classification of the infants sleep states: active (AS) and quiet sleep (QS). **(B)** Sensor-level signals were filtered into narrow frequency bands and cortical activity signals were extracted by source reconstruction into 58 cortical areas (parcels) that covered all four cortical regions (F, frontal; C, central; T, temporal; and O; occipital) using an infant head model. *a.u.* stands for arbitrary units. **(C)** Functional connectivity between cortical parcels was computed as phase-phase correlations (PPCs), yielding frequency- and sleep state–specific connectivity matrices (networks) in each subject. These networks were statistically compared between groups to find the treatment effects. **(D)** The replicability of the FNI-affected networks was assessed by comparing their spatial distributions, relation to HCs, and development over time in RCT1 and RCT2.

### Subjects

This study utilized data from preterm infants born between 26 to 34 weeks of gestational age (GA) collected in a parallel-group randomized controlled trial (RCT1, N=137) and a multisite replication trial (RCT2; N=135). RCT1 (registered at ClinicalTrials.gov, #NCT01439269) was conducted at a level IV NICU at Morgan Stanley Children’s Hospital of New York, Columbia University Medical Center (CUMC). RCT2 (registered at ClinicalTrials.gov, #NCT02710474) was performed at two level IV NICUs: the Morgan Stanley Children’s Hospital of New York, Columbia University Medical Center (RCT2-NY), and the University of Texas Health’s affiliate University Health System, San Antonio (RCT2-SA). The study design was approved by the CUMC Institutional Review Board and informed consent was obtained from a parent or guardian for each subject. The study, randomization procedures, inclusion/exclusion criteria, and the baseline characteristics of the cohorts have been described previously ^36,37^.

The analyses also include a group of term-born HC infants (N = 66) born at 40.4 ± 1.1 weeks GA (mean ± SD) from the Children’s Hospital of Helsinki University Central Hospital. These data were gathered from study groups of previously published cohorts ^5,9,47,48^. The recruitment, consent, and study procedures were approved by the Ethics Committee of the Helsinki University Central Hospital.

### Intervention protocol

The mother-infant dyads were assigned to either SC or FNI treatment by a permuted block randomization described previously ^26,37^. The SC cohort received established routine care of the NICU provided by health care professionals. The FNI cohort received SC plus FNI. FNI is a NICU-based multimodal intervention, which aims to facilitate the development of the emotional parent-infant connection during the NICU stay. A detailed description can be found in ^26^. Briefly, the FNI activities were facilitated by nurture specialists during 1-hour calming sessions, beginning at the earliest possible time point after delivery, which was 1 week on average. The initial activities included firm sustained touch, maintained eye contact, exchange of scent cloths, and deep emotional vocal soothing. When the infant was medically stable, nurture specialists commenced the facilitation of clothed, or skin-to-skin care paired with deep emotional expression. Nurture sessions were performed 3.5 times per week on average over the hospitalization period.

### EEG acquisition

In RCT1 and RCT2, longitudinal high-density EEG was acquired at several timepoints (N = 1-4) during daytime sleep between 33 to 45 weeks of conceptional age (CA) with a 128electrode system (EGI Magstim, Spring Gardens, Whitland, SA32 0HR, UK) using a vertex reference and 1 kHz sampling frequency. In the HC cohort, EEG was collected at term age (41.4 ± 1.4 weeks CA) using 19-channel caps (ANT-Neuro, Berlin, Germany) and a NicOne EEG amplifier (Cardinal Healthcare, Ohio/Natus, Pleasanton, USA) using a vertex reference and 256 Hz or 500 Hz sampling frequency. The recording details can be found in ^36,37,49^.

The EEG recordings were classified into epochs of active sleep (AS) and quiet sleep (QS) by experts (SV and PY) based on standard criteria ^50^. Briefly, AS is characterized by continuous oscillations, transient eye movements, and possible muscle activity in frontal or temporal regions. QS is determined by the presence of a clear tracé discontinue/alternante with prominent spontaneous activity bursts, regular breathing and possibly reduced muscle tone. AS or QS recordings with under 3 (RCT2) or 5 minutes (RCT1) of good quality data were rejected. Due to the absence of recordings between 37-39 weeks CA in the replication cohorts, the age range around term was rendered from the original 37-43 weeks CA to 39-43 weeks CA. A reanalysis of the results from the original cohort (RCT1) within this age range showed comparable results (Supplementary Fig. 1). The developmental analysis utilized the full set of recordings from 33 to 45 weeks CA. The final sample sizes of good-quality recordings are presented in Table 1.

**Table 1.** Sample sizes of good-quality EEG recordings in all cohorts.

| Cohort | All recordings<br>(33-45 weeks CA) |  | Term age recordings<br>(39-43 weeks CA) |  |
| --- | --- | --- | --- | --- |
|  | AS | QS | AS | QS |
| RCT1 | FNI: 170<br>SC: 131 | FNI: 168<br>SC: 128 | FNI: 41<br>SC: 35 | FNI: 38<br>SC: 33 |
| RCT2-NY | FNI: 60<br>SC: 59 | FNI: 61<br>SC: 63 | FNI: 28<br>SC: 25 | FNI: 29<br>SC: 29 |
| RCT2-SA | FNI: 37<br>SC: 40 | FNI: 38<br>SC: 37 | FNI: 15<br>SC: 18 | FNI: 16<br>SC: 16 |
| HC | - | - | HC: 48 | HC: 57 |

### EEG preprocessing

The data were imported to Matlab R2024b (MathWorks, Natick, MA) for preprocessing and analysis, using the same pipeline as in the original study ^38^. The direct current (DC) was removed by subtracting the mean from all epochs followed by filtering from 0.4 to 40 Hz (5th order high- and low-pass Butterworth filters) and down-sampling to 100 Hz. In RCT1 and RCT2, channels 125 to 128 were excluded due to location on the face. All channels were visually inspected and bad channels with prominent artefacts, such as loose or unstable electrodes, electrical interference, flat signals, and excessive muscle activity or movement artifacts, were excluded. In addition, time periods with prominent artefacts in several channels were excluded. The electrocardiographic artefact (ECG) was removed from the RCT1 and RCT2 cohorts using independent component analysis (ICA) ^51,52^ and automatic detection of the ECG-related components based on their temporal structure ^53^ confirmed manually by a reviewer (PY). Finally, the data were converted to average montage and filtered into 24 frequency bands of interest using the same bank of filters as reported previously ^38^. Briefly, these Butterworth filters have been designed to span the range with central frequencies (Fc) from 0.5 to 33 Hz at semi-equal distances on a logarithmic scale with passband frequencies computed as 0.85×Fc and 1.15×Fc and stop-band frequencies as 0.5×Fc and 1.5×Fc.

### Computation of cortical signals and functional connectivity

The preprocessed and filtered sensor-level signals were next reconstructed to source level using an infant head model ^5^ with tissue conductivities as 0.43, 0.2, and 1.79 S/m for the scalp, skull, and intracranial volume, respectively. The inverse solution was computed using dynamic statistical parametric mapping ^54^ into a source space with N = 8014 normally oriented cortical dipoles. The dipoles were collated into N = 58 cortical parcels by K-means clustering, and parcel signals were computed as weighted average of the corresponding source signals.

Functional connectivity was assessed from source reconstructed cortical signals by computing pairwise interactions between all parcels with PPCs, using the debiased weighted phase-lag index ^55^. This yielded individual functional connectivity matrices of size 58 x 58 for each frequency band and sleep state. As in the original study ^38^, infant age at recording was regressed from the PPC value of each connection with linear fitting, which yielded the studentized residual as an age-corrected measure of connectivity. Age-corrected values were used to construct the FNI-affected cortical networks.

### Replication strategy of FNI-affected cortical networks between trials

To construct FNI-affected cortical networks, we first performed a systematic comparison of PPC strength between the FNI and SC cohorts (one-tailed Wilcoxon rank-sum tests, alpha = 0.05) in all 1653 connections for both directions (FNI > SC and FNI < SC), as was done in our original work ^38^, for RCT1, RCT2-NY and RCT2-SA. Here, the goal was to assess whether cortical activity networks can be found at the same frequencies as identified in the original study: 0.5 Hz, 3.1 Hz, 5.3 Hz, and 11.1 Hz in AS and 1.8 Hz, 3.1 Hz, 5.3 Hz, and 11.1 Hz in QS. We first computed the proportion of connections with significant group differences (*K*) on the full frequency range to assess the overall spectral similarity of findings and then analyzed specifically the *a priori* defined frequencies. The potential rate of false positives was identified using the adaptive positive false discovery rate (FDR) ^5,56^.

### Criteria for successful replication

The criteria for successful replication was if the *a priori* defined frequencies showed a network whose extent surpassed the FDR threshold (q = 0.05) in the same direction (FNI < SC vs. FNI > SC) as in the original study.

### Statistical analysis

The spatial properties of the constructed FNI-affected networks were next compared between RCT1 and RCT2 trials. Here, trial-specific networks were aggregated to regional level (for details, see Supplementary Material) and the resulting regional matrices were compared across trials with the Mantel test (Spearman correlation, 5000 iterations) ^57^. We applied *post hoc* correction for multiple comparisons with the Benjamini-Hochberg procedure for each sleep state separately ^58^.

Next, PPC strength in the trial-specific FNI-affected networks was compared between FNI vs. HC and SC vs. HC (two-tailed Wilcoxon rank sum tests) in both trials similarly to the original study ^38^. While previously we investigated only alpha frequency networks (11 Hz), here we opted to analyze all the predefined frequencies of interest. Due to uncontrollable variance in the fetal age determination between health care practices (USA vs Finland), age correction was not applied in this analysis, in line with the original paper. Effect size was estimated using the rank biserial and multiple comparisons correction was computed *post hoc* with the Benjamini-Hochberg correction separately for each sleep state ^59^. Outliers exceeding 3 x standard deviation (SD) were rejected from each cohort.

Finally, we performed a developmental analysis on FNI-affected networks in both trials, which extends the scope of the original study. We used the EEG recordings from 33 to 45 weeks CA, with 3 recordings rejected due to recording outside this time window. First, we compared PPC strength within the group difference networks defined earlier between FNI and SC groups at preterm age, defined as 33 to 37 weeks CA (two-tailed Wilcoxon rank sum test). Next, we computed Spearman correlation of mean PPC in the group difference networks as a function of CA in both groups and trials. The resulting correlation coefficients were compared between the groups using *cocor* (independent groups two-tailed test) ^60^. Multiple comparisons were controlled *post hoc* with the Benjamini-Hochberg procedure. To ensure that the repeated measures data did not bias the correlations, we checked the correlations also using a linear mixed-effects model with CA as the fixed effect and subject ID as a random effect (Supplementary File 1). All statistical analysis results are reported in Supplementary File 1.

### Analysis software

Data preprocessing and analyses were performed with Matlab R2024b (MathWorks, Natick, MA). ICA was implemented using the Picard package ^51,52^. Source reconstruction was computed with Brainstorm ^61^ and OpenMEEG ^62^. Benjamini-Hochberg multiple comparisons corrections were computed with the SDM implementation ^59^. Comparison of correlation coefficients was computed with the *cocor* tool ^60^. Brain networks were visualized using BrainNet Viewer ^63^. Implementation of network analysis for group differences is provided as MATLAB code at https://github.com/pauliina-yrjola/Preterm-Phase.

## Results

We computed FNI-affected networks in each trial, followed by comparing spatio-spectral properties of these networks across trials. Following our previous work, the neurodevelopmental effect was assessed by comparing FNI and SC connectivity strength against the same independent group of healthy controls (HC). Finally, the original analysis was extended to investigate the development of the FNI-induced networks from early preterm to term age.

### FNI effects on cortical networks replicate in independent trials

The replication cohort RCT2-NY showed FNI-related networks that compared well with those observed in RCT1 (Fig. 2). The key FNI-effects in both RCT1 and RCT2-NY included: broadband decrease in connectivity strength in AS with peaks at high-delta (3.1 Hz, *p* < 0.05, *K* = 11 % in RCT1 and *K* = 12 % in RCT2-NY) and alpha (11 Hz, *p* < 0.05, *K* = 8.3 % in RCT1 and *K* = 8.0 % in RCT2-NY). A similar decrease was seen somewhat less prominently in QS (peaks at 3.1 Hz, 5.3 Hz and 11 Hz, *p* < 0.05, *K* up to 7.6 % in RCT1 and 6.7 % in RCT2-NY). Additionally, the clear peak of stronger PPC in FNI found at low-delta frequencies of QS in RCT1 was present also in RCT2-NY (*p* < 0.05, *K* = 13 % in RCT1 and *K* = 8.2 % in RCT2-NY). Based on our criteria for a successful replication, (section “Criteria for successful replication” in Methods), we conclude that the key findings replicate well between study cohorts carried out in the same center at different times (RCT1 and RCT2NY).

**Fig. 2.**
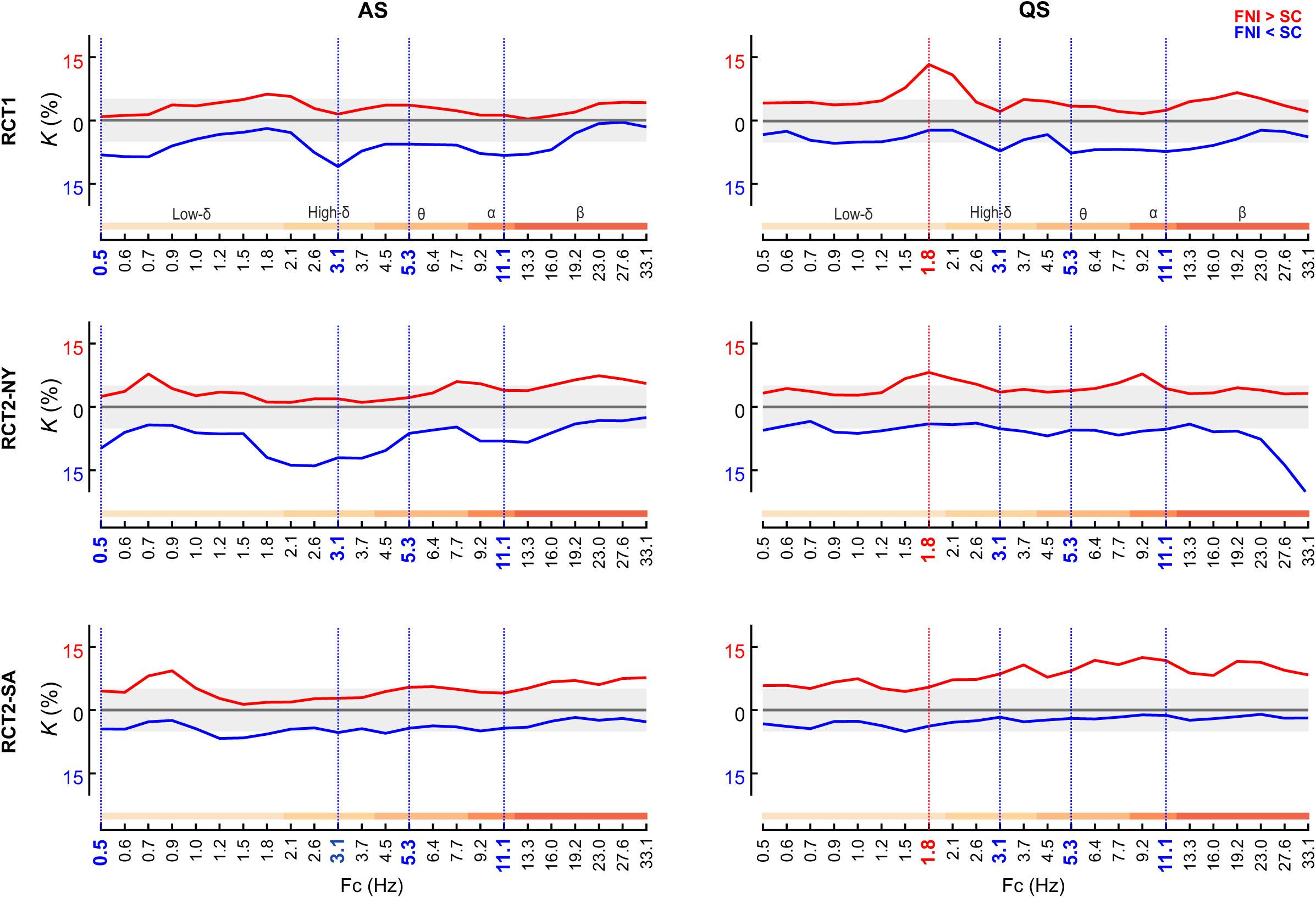
A spectral comparison of FNI effects on cortical activity networks in different RCT cohorts. The proportion *K* (%) of connections showing a significant difference between FNI and SC groups (two one-tailed Wilcoxon rank sum tests) in age-corrected PPC strength in both the earlier trial (RCT1) and the replication trials (RCT2-NY and RCT2-SA). Red lines depict networks of increased (FNI > SC) and blue networks of decreased (FNI < SC) connectivity strength in the FNI infants as compared to the SC infants in AS (left) and QS (right). Vertical dashed lines highlight the frequencies of interest fixed as those identified in the original study ^38^. The grey shaded area depicts the level of potential false positives.

In the external validation cohort, RCT2-SA, previous findings were replicated to a somewhat lesser extent at high-delta in AS (3.1 Hz, *p* < 0.05, *K* = 5.3 %) and low-delta in QS (1.8 Hz, *p* < 0.05, *K* = 5.3 %). The other networks of interest did not reach statistical significance that surpassed the FDR level (for possible reasons, see Discussion).

### The spatial distribution of FNI-affected cortical networks is similar across trials

To assess whether the cortical activity networks are spatially similar between the trials, we compared their spatial distributions on a regional level (Fig. 3). In AS, the most prominent networks, high-delta, were comparable between RCT1 and RCT2-NY (3.1 Hz, *p* = 0.032, *p*_BH = 0.13, rho = 0.58). Moreover, in QS, theta networks showed significant comparability (5.3 Hz, *p* = 0.043, *p_*BH = 0.09, rho = 0.44) and low-delta and alpha networks modest comparability (*p* = 0.064, *p_*BH = 0.09, rho = 0.62 and *p* = 0.066, *p_*BH = 0.09, rho = 0.39, respectively) between trials. Taken together, despite the inherently high biological and analytical variability in spatially resolved network details, there is a striking match across trials, especially in the QS state.

**Fig. 3.**
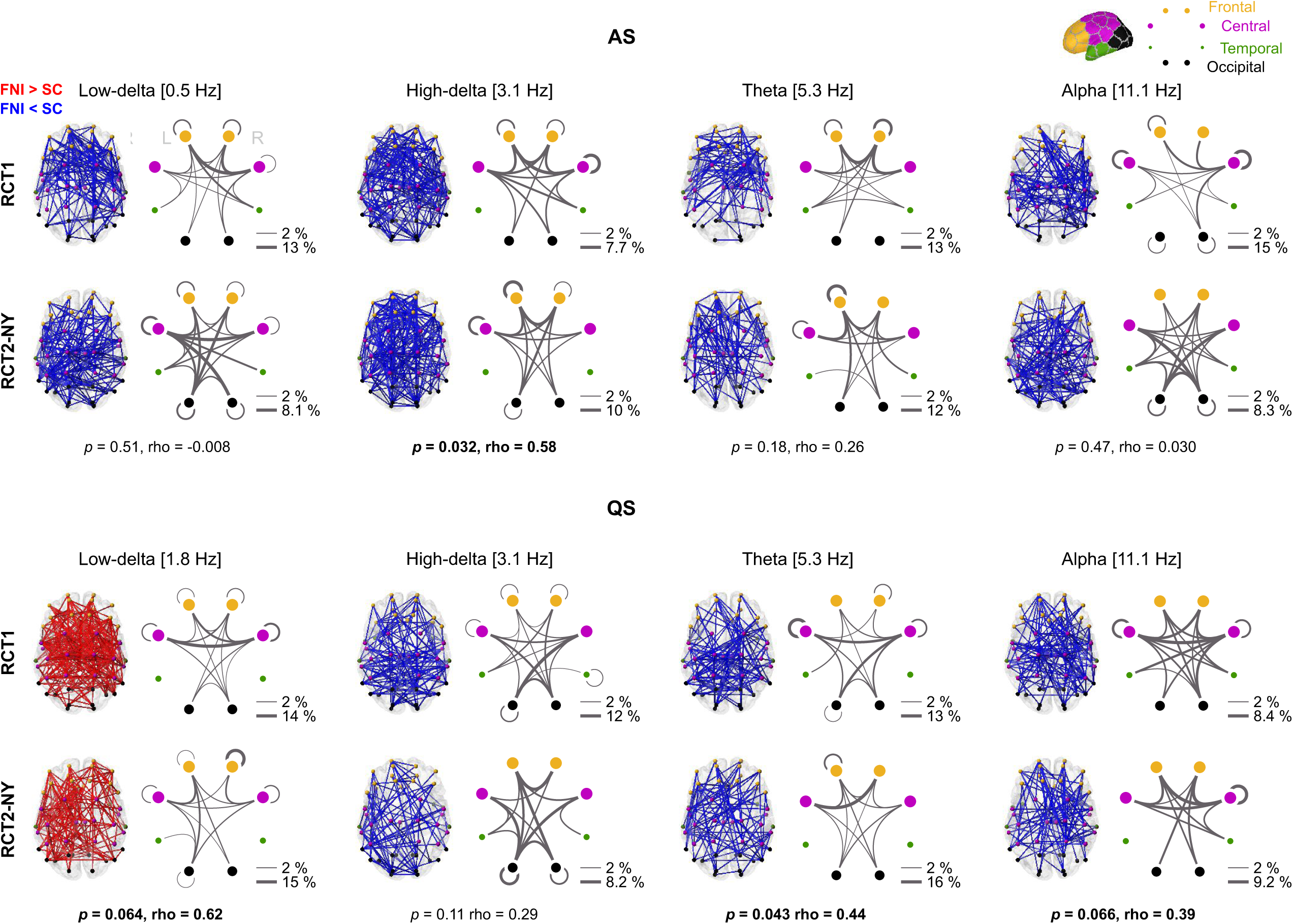
Comparison of the spatial distribution of FNI-affected cortical networks. The topologies of the group difference networks are shown on a cortical surface across all frequencies of interest and for both RCT1 and RCT2-NY. Red lines depict networks of increased (FNI > SC) and blue networks of decreased (FNI < SC) connectivity strength in the FNI infants as compared to the SC infants in AS (top) and QS (bottom). Spider plots represent the involvement of regions in the group difference networks: frontal (orange), central (magenta), temporal (green), and occipital (black). Line widths indicate the proportion (%) of connections between each region pair normalized by network size, and node sizes indicate the share of parcels in each region. The *p*- and rho-values of a Mantel test indicating the similarity of RCT2-NY networks to RCT1 at the level of regions are shown below the plots.

In RCT2-SA (Supplementary Fig. 2), significant comparability to RCT1 networks was found at low and high-delta frequencies in AS (0.5 Hz, *p* = 0.011, *p_*BH = 0.044, rho = 0.54 and 3.1 Hz, *p* = 0.036, *p_*BH = 0.072, rho = 0.48) and at low-delta in QS (1.8 Hz, *p* = 0.085, *p_*BH = 0.34, rho = 0.34).

### FNI renders cortical networks of preterm infants closer to term-born controls

Next, we attempted to replicate the previously found comparability of cortical activity networks in the FNI cohort compared to term-born HCs ^38^ extending the previously performed analysis of alpha frequencies to all the frequencies of interest. Similarly to RCT1, in RCT2-NY, we found that the mean connectivity strengths of FNI infants did not differ significantly from HCs during either sleep state (Fig. 4; *p* > 0.11, *p*_BH > 0.16, rank-biserial r = 0.02-0.22) in all frequences except low-delta in QS, where connectivity was significantly higher in FNI compared to HC (*p* < 0.001, *p*_BH < 0.001, rank biserial r = 0.68). In contrast, SC infants showed significantly stronger connectivity compared to HC at the same frequencies (*p* < 0.001, *p*_BH < 0.001, rank-biserial r = 0.63-0.85). RCT2-SA (Supplementary Fig. 3) showed essentially similar results in all frequencies with slightly more variation in FNI connectivity strengths compared to HC.

**Fig. 4.**
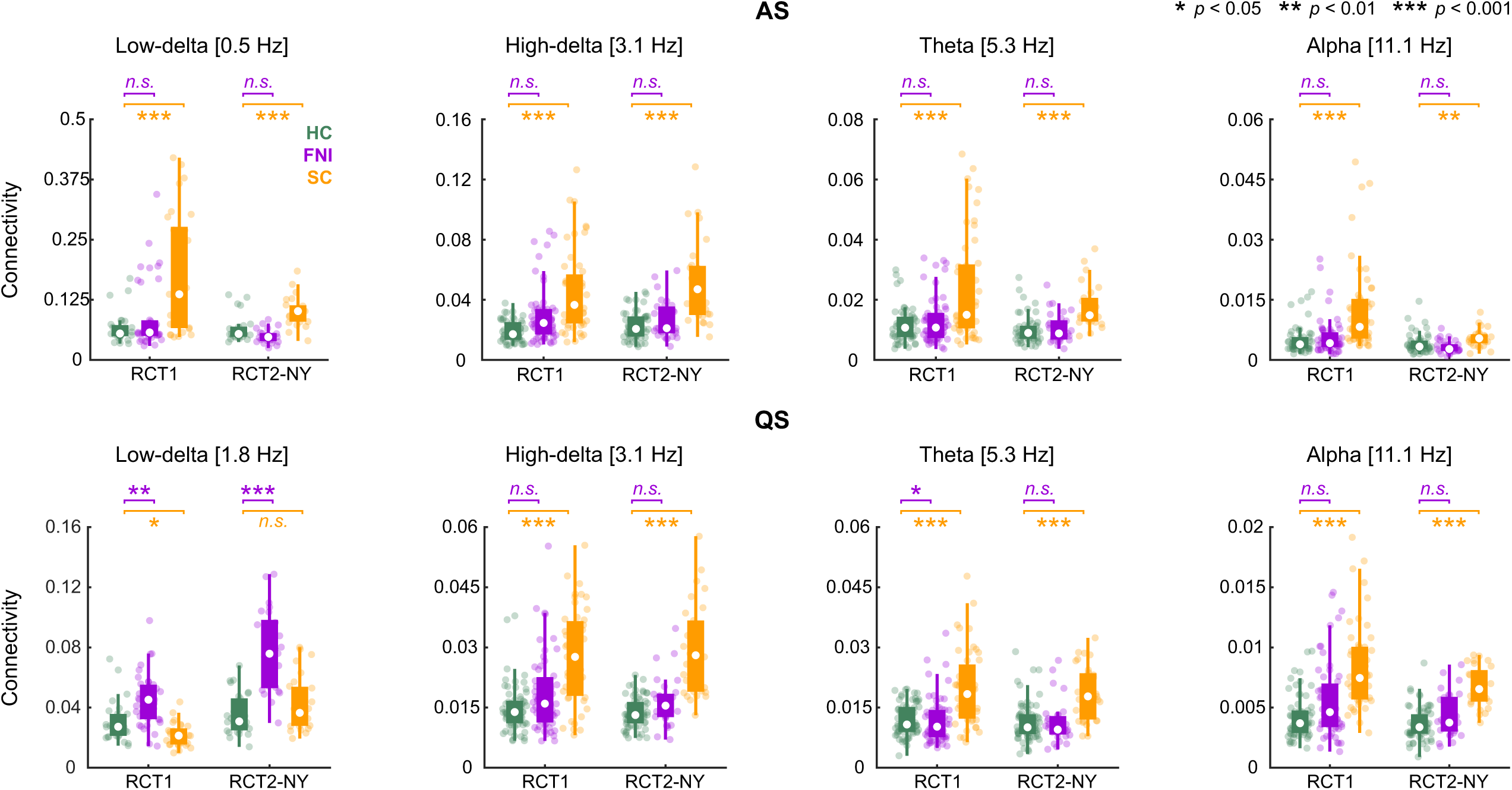
Comparison of the FNI-affected network strength in the preterm (FNI, SC) and term-born (HC) infants. Boxplots depict mean network strength at term age in the HC (green), FNI (purple) and SC (orange) groups for the RCT1 and RCT2-NY cohorts in AS (top) and QS (bottom). White dots depict the group median, the lower and higher edges of the box indicate the interquartile range, and the whiskers indicate the most extreme data points not considered outliers. Scatter points indicate individual mean network strength. Asterisks denote significant group difference (two-tailed Wilcoxon rank-sum test) for FNI vs HC (purple) and SC vs HC (orange). *n.s*. stands for non-significant.

### Cortical network trajectories of FNI and SC infants diverge during the neonatal period

Finally, we extended our analysis to investigate the timing of the observed network differences throughout the period from 33 to 45 weeks of CA (Fig. 5). We found that at early preterm age (33 to 37 weeks CA), FNI and SC networks do not differ significantly at any frequency (*p* > 0.07, *p*_BH > 0.13, rank-biserial r = 0.007-0.27), but after that the trajectories diverge. The main effect was a replicable age-related decrease in the network strength of FNI infants at several frequencies; in AS, a replicable decrease was present in the theta band (*p* < 0.001, *p*_BH < 0.01, rho up to -0.51), while in QS, the decrease was replicable over high-delta, theta and alpha frequencies (*p* < 0.01, *p*_BH < 0.03, rho up to -0.55). In contrast, no such consistent changes were observed in the SC infants. The divergence of trajectories in these frequencies was confirmed by a comparison of correlation coefficients between groups, which revealed significant differences in network development (*p_*cocor < 0.01, *p_*cocor_BH < 0.02, Fisher z up to 3.4). Similar findings were present in RCT2-SA (Supplementary Fig. 4).

**Fig. 5.**
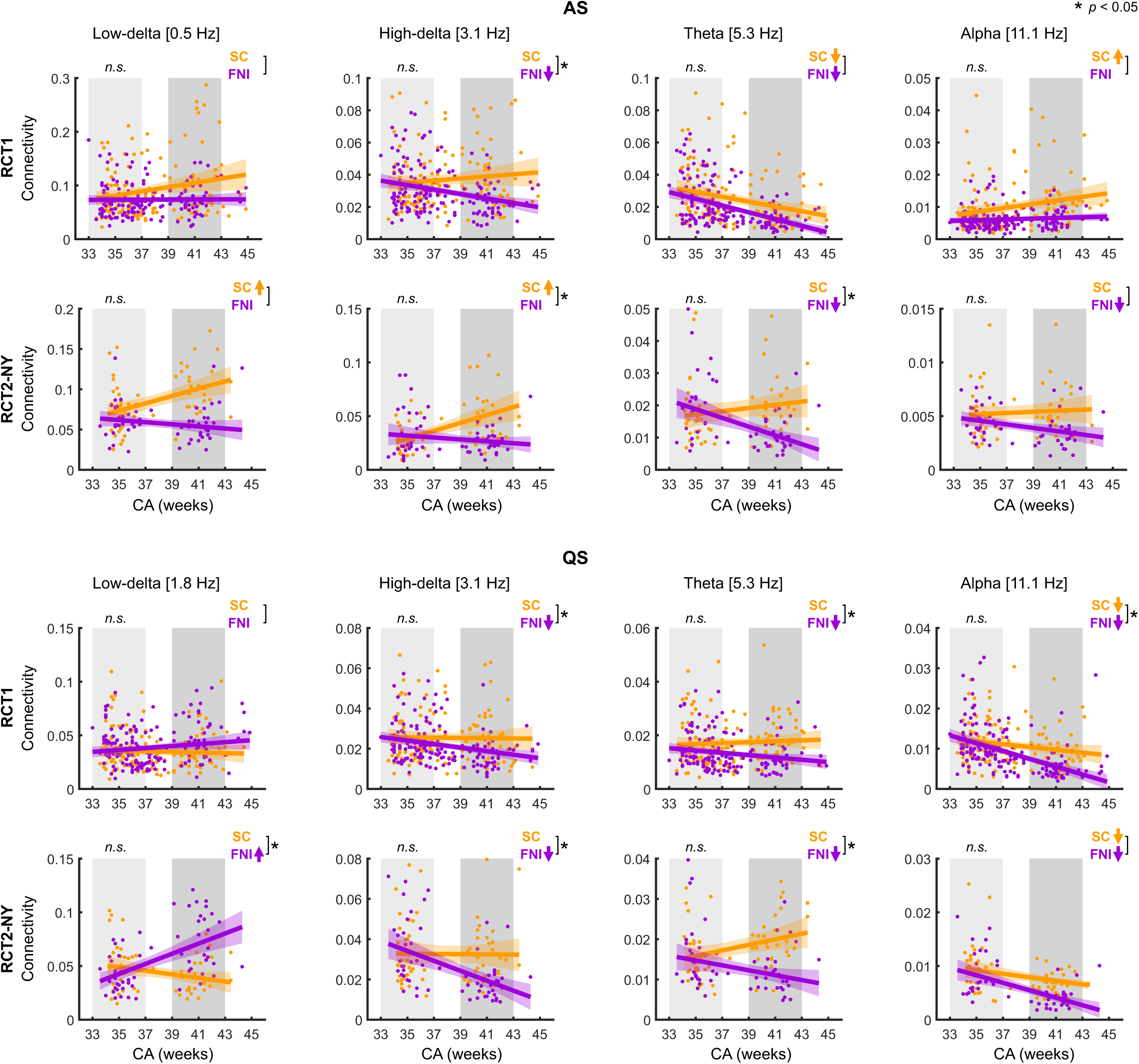
Development of FNI-affected networks from 33 to 45 weeks CA. Mean network strength is presented as a function of CA in the FNI (purple) and SC (orange) groups for the RCT1 and RCT2-NY cohorts in AS (top) and QS (bottom) with confidence intervals shaded in the same color. Grey shaded areas depict the early preterm age (light grey) and term age (dark grey) windows. Group difference (two-tailed Wilcoxon rank-sum test) was computed between FNI and SC groups in the preterm age window. *n.s.* stands for non-significant results after Benjamini-Hochberg correction. Correlation of network strength with age (Spearman correlation) was computed for both groups; next to each plot, arrows indicate significant (*p* < 0.05) increase (upwards) or decrease (downwards) of PPC strength with CA in each group. Asterisks next to the square bracket indicate a significant (*p* < 0.05) difference in the correlation coefficients of the groups (two-tailed independent groups comparison of correlations, *cocor*).

## Discussion

Our present findings show that the effects of FNI on the early developing cortical networks replicate robustly in an independent trial, particularly within the same research site. Moreover, we show that the development of FNI-affected networks diverges from the infants receiving standard care, suggesting neurodevelopmental effects as early as during the NICU period. These results together support the hypothesis that FNI modulates cortical networks across early maturation and mitigates the effects of prematurity. The results of the study fit well with the existing notion that EE interventions administered in the NICU may be beneficial for early brain development after preterm birth ^19^ as well as into the general framework of early development of cortical activity networks ^64^. To our knowledge, the present study is the first to show the replicability of improvements on the development of neuronal activity networks when applying a NICU-based EE intervention.

Several lines of recent evidence point to the fact that newborn brain maturation is characterized by context- and frequency-dependent changes in the coherent activity of large-scale brain networks ^9,38,64–66^, which are predominantly directed towards an age-related decrease in synchrony ^64,65^. Our present findings show that FNI affects cortical networks most prominently and reproducibly in fronto-central areas but involving connections throughout the brain with small cohort-specific differences. The most salient effect is the observed broadband decrease of PPC strength, involving mostly theta and alpha frequencies. This is in line with previous reports, which have shown similar FNI-affected decreased strengths of coherence and phase synchrony in fronto-central areas rendering FNI infants closer to term-born controls than infants receiving SC ^38,67^. This decreased connectivity found repeatedly in FNI infants across trials can be linked to several frameworks. Developmentally, preterm brain maturation is characterized by a transition from highly synchronous activity bursting (a.k.a. spontaneous activity transients, SATs) to desynchronized continuous activity which overcomes bursting at around term age ^64,68^. The decreased connectivity strength of FNI could thus reflect a more mature level of early neuronal communication. Neurocognitively, the decreased strengths of neuronal synchrony have previously been associated with several benefits, such as improved neurocognitive outcomes in later toddlerhood ^38^ and better parent-infant interaction dynamics ^69^. This may reflect the involvement of these networks in the emerging higher cognitive abilities observed at these frequencies in adults ^70–72^. An interesting exception to the general decrease in connectivity is the low-delta frequency, particularly in QS, which has previously been found to increase with age ^64^ and to correlate positively with neurocognitive performance in preterms ^6^. Strikingly, FNI introduces a replicable increase in connectivity at this frequency, highlighting mutually coexisting multiplex networks that brain function relies on ^71,73^.

While the replication conducted at the same site (RCT2-NY) robustly reproduced the previous results, the results in a validation cohort recorded at an external site (RCT2-SA) were somewhat less clear, with several frequency bands showing no group difference surpassing the FDR level or networks of the opposite direction. Assessing these observations calls for careful consideration of social, technical, medical care related differences between sites. For instance, demographic and social differences between New York and rural San Antonio, as well as increased awareness and between-center differences in the integration of parents in their infants’ care at the NICU may provide plausible explanations to the observed unclarity of generalization. In addition, it has been shown that cortical networks reflect a high level of brain organization ^40–42^ large variability in preterm infants ^47,74^. This makes group-level results very susceptible to even minor changes within the cohort, particularly with small sample sizes such as those of RCT2-SA.

The neurodevelopmental alterations on functional cortical networks caused by FNI call for a reasonable mechanistic explanation; however, due to its multimodal nature, the investigation of direct causal links between FNI and neuronal activity is challenging. Here we showed that the cortical activity networks of the intervention groups are comparable at preterm age (here defined as 33-37 weeks CA) but diverge after, indicating that FNI modulates networks to comparable strengths of connectivity relative to term born controls at term age. Neurobiologically, preterm birth inflicts a series of dysmaturational events, with or without definite white matter injury ^1,2^, leading to decreased density of the core structural connections ^46^. At term age, alterations in functional networks ^5,6^, large variability in functional networks between individuals ^47^, and an increased association of functional network strength to neurodevelopmental outcome ^6^ are observed in preterm infants compared to their term-born counterparts, suggesting that preterm infants make the most out of their remaining injured network. Behaviorally, FNI is set apart from other EE interventions, which focus mostly on external sensory experiences of human voice, music or massage therapy ^22,23,25^, by emphasizing the importance of the deep emotional engagement between the mother-infant dyad. While sensory stimuli are undoubtedly produced, FNI effects are most likely mediated via the physiological stabilization of the infant due to recruitment of the endogenous parent-infant behaviors. This may support the restoration of white matter injury or neuronal maturation underlying myelination, a key conditioner of neuronal networks ^75^.

The limitations of the present study may include increased awareness of the FNI intervention. Most importantly, the SC practices have changed over the decade between the trials (RCT1 commencing in 2008 and RCT2 in 2017) with new and beneficial practices, such as unrestricted visiting hours, family centered care and promotion of early breastfeeding, adopted in many NICUs, with clear differences between clinical centers. Thus, the adoption of similar nurturing practices to mothers of the SC group cannot be ruled out. Additionally, we were unable to investigate the previously found association of lower PPC strength with neurocognitive outcomes due to the loss of follow-up measurements as a consequence of COVID. However, we believe that the set of EEG recordings from the cohort obtained at the same site, CUMC in New York, is sufficient to allow the conclusions made about the main effect, a replicable decrease of PPC strength in infants receiving FNI, and the extension of the divergence in trajectories across the neonatal period.

In conclusion, the present work shows that the FNI effects on functional brain networks of preterm infants were replicated robustly in an independent trial. Moreover, we showed that the six weeks of FNI between preterm and term age lead the infants to a different trajectory of brain development compared to SC, resulting in comparable network strength to healthy term-born counterparts at term age. The findings together provide evidence which point to the fact that encouraging parent-infant contact and emotional connection during the NICU stay can be beneficial in restoring early neurodevelopment in preterm infants. This evidence may be leveraged to support the use of FNI, whose simplicity, cost-effectiveness, and global accessibility render its global potential substantial.

## Acknowledgements

We thank parents and their infants for participation, as well as the research nurses involved in running the intervention trial and contributing to the technical assistance in both the FNI cohort and the healthy term cohort.

## Author contributions

Conceptualization and coordination of interventions and related data collection: MGW, MMM

Coordination of data collection of the healthy term cohort: SV

Conceptualization of the EEG analyses: PY, AT, SV

Classification of infant sleep states: SV, PY

Performance of the EEG analyses: PY

Contribution of analytical tools: AT

Visualization of results: PY

Writing—original draft: PY

Writing—review & editing: SV, AT, MGW, MMM

## Consent statement

Informed consent was obtained from a parent or guardian for each subject.

## Data and materials availability

All data needed to evaluate the paper’s conclusions are provided in the paper and supplementary materials. MATLAB scripts of the network analysis for group differences are available in https://github.com/pauliina-yrjola/Preterm-Phase.

