## Supplementary Material for "Early development of cortical networks is modulated by Family Nurture Intervention: a multicenter replication study"

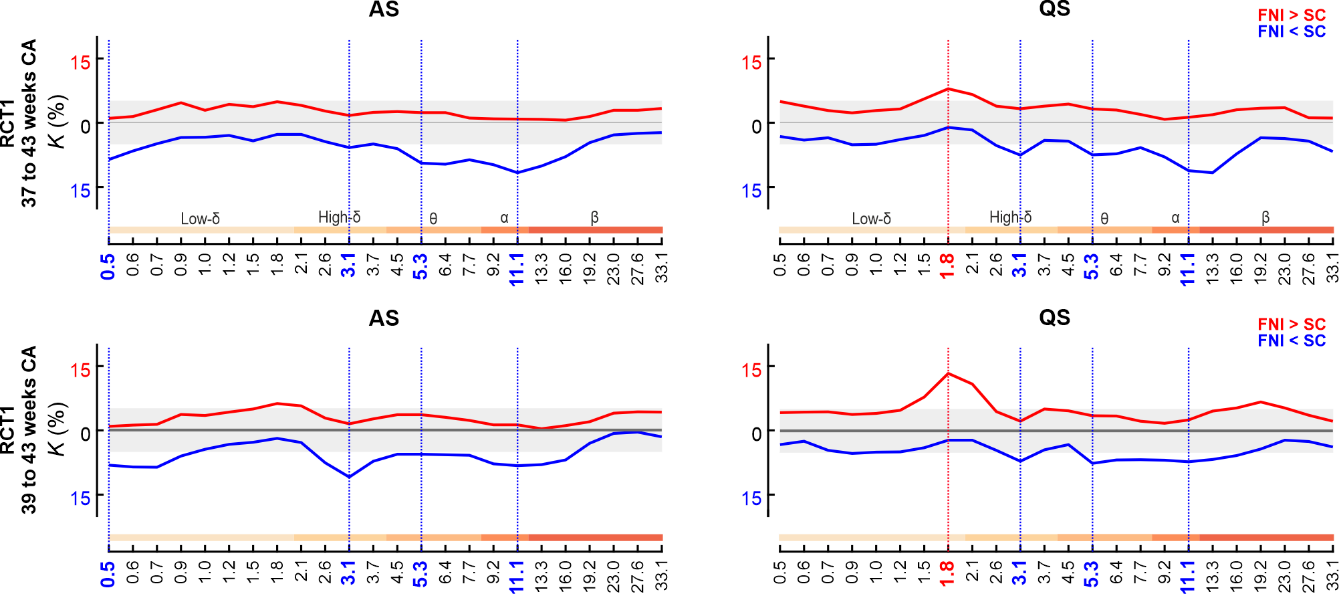

**Supplementary Fig. 1. A comparison of FNI effects on cortical activity networks in RCT1 in two age windows.** The proportion *K* (%) of connections showing a significant difference (two one-tailed Wilcoxon rank sum tests) in age-corrected PPC strength between treatment groups (FNI and SC) in RCT1 in a broad (37 to 43 weeks CA) and narrow age range (39 to 42 weeks CA). Red lines depict networks of increased and blue networks decreased connectivity strength in the FNI infants as compared to the SC infants in AS (left) and QS (right). Vertical dashed lines highlight the frequencies of interest fixed as those identified in the original study (broad age range). The results are retained in the narrower age range with comparable peak frequencies.

**Regional aggregation of cortical networks for spatial comparison**

To enable the comparison of network spatial distributions while accounting for the effect of small differences in individual anatomy and/or cap positioning, the spatial information was aggregated from parcel level to regional level. Here, the number of parcel-level connections between each pair of regions (frontal, central, temporal, and occipital) were summed and normalized per number of possible connections between the respective region pair. This procedure was performed for inter- and intrahemispheric connections yielding regional matrices of size 8 x 8, which were used in statistical analysis.

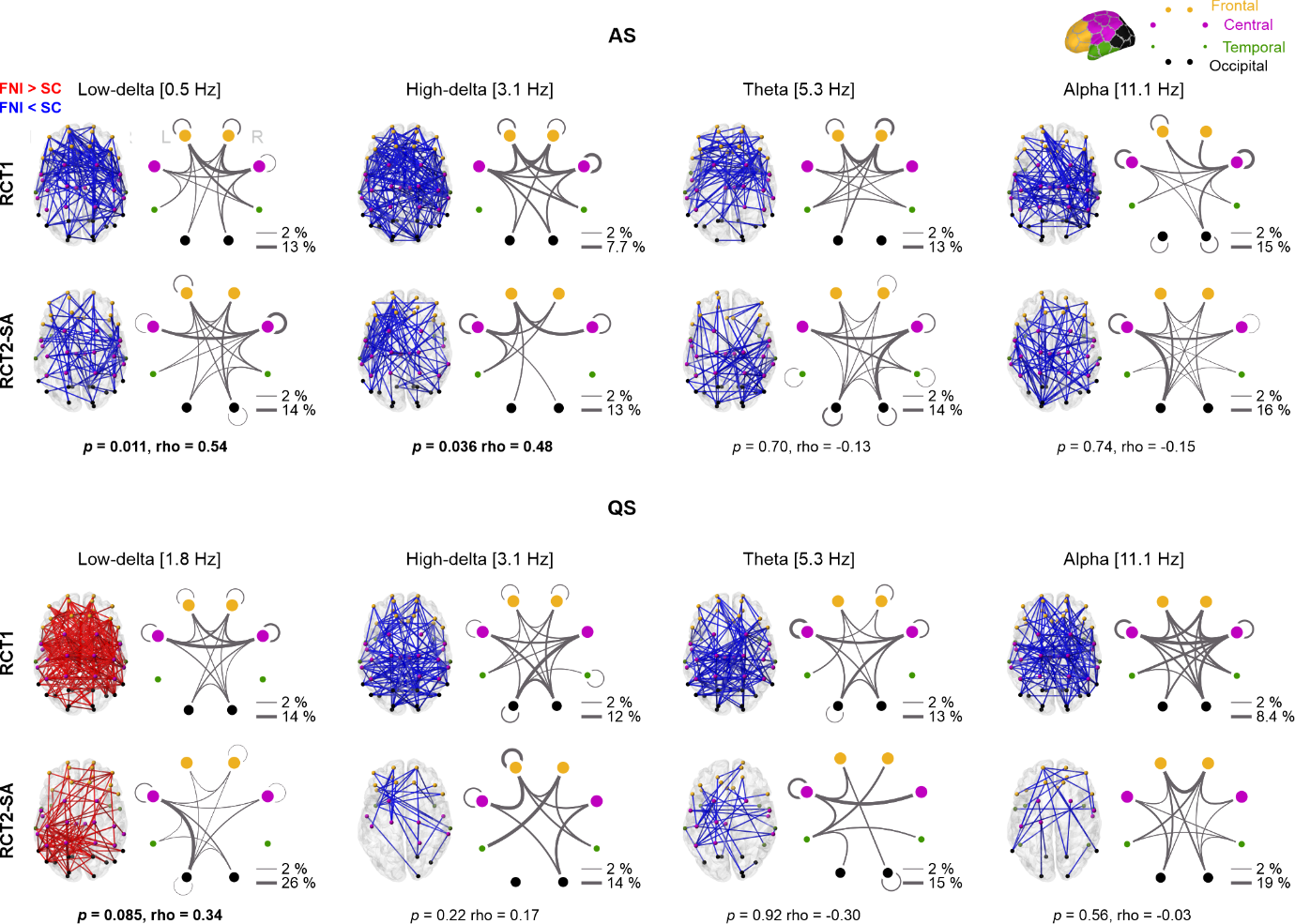

**Supplementary Fig. 2. Comparison of the spatial distribution of FNI-affected cortical networks between RCT1 and RCT2-SA.** The topologies of the group difference networks are shown on a cortical surface across all frequencies of interest and for both RCT1 and RCT2-SA. Blue lines indicate connections with decreased and red connections with increased connectivity strength in the FNI infants as compared to SC infants in AS (top) and QS (bottom). Spider plots represent the involvement of regions in the group difference networks: frontal (orange), central (magenta), temporal (green), and occipital (black). Line widths indicate the proportion (%) of connections between each region pair normalized by network size, and node sizes indicate the share of parcels in each region. The *p*- and rho-values of a Mantel test indicating the similarity of RCT2-SA networks to RCT1 at the level of regions are shown below the plots.

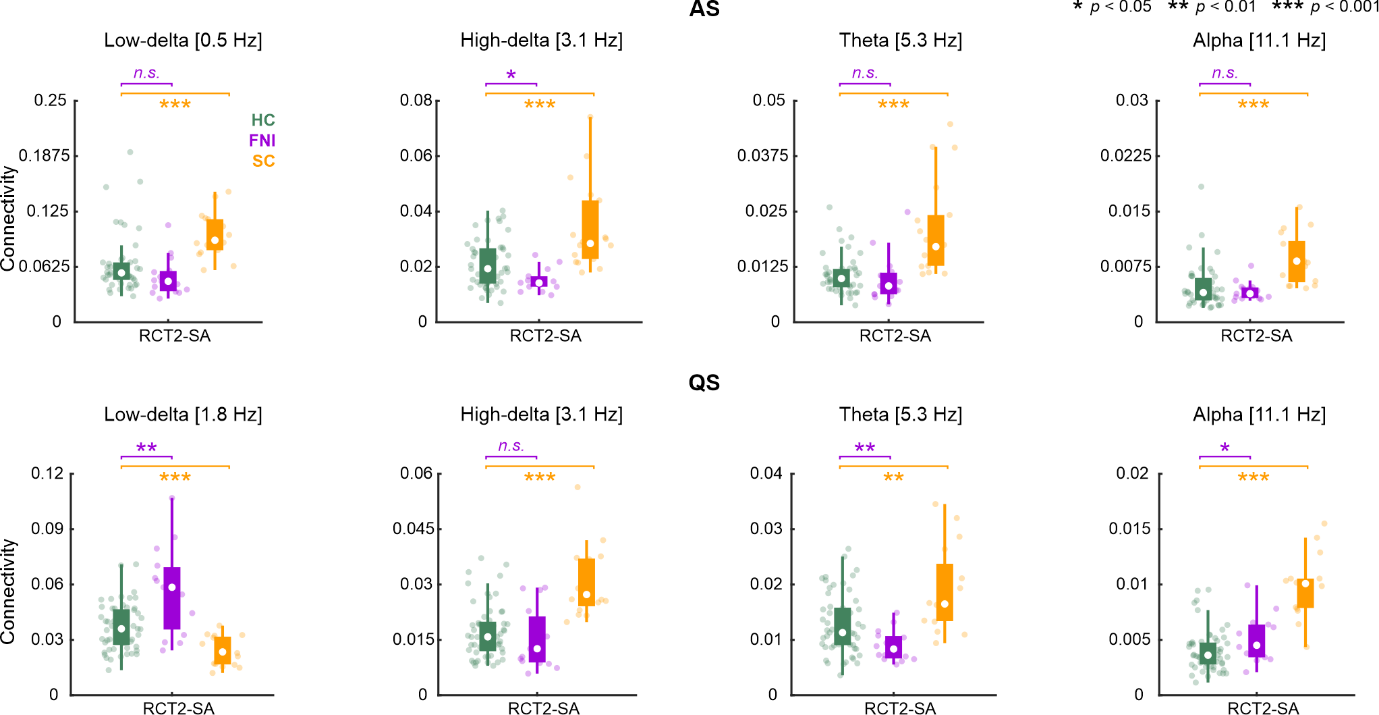

**Supplementary Fig. 3. Comparison of the FNI-affected network strength in the preterm (FNI, SC) and term-born (HC) infants in RCT2-SA.** Boxplots depict mean network strength at term age in the HC (green), FNI (purple) and SC (orange) groups for the RCT2-SA cohort in AS (top) and QS (bottom). White dots depict the group median, the lower and higher edges of the box indicate the interquartile range, and the whiskers indicate the most extreme data points not considered outliers. Scatter points indicate individual mean network strength. Asterisks denote significant group difference (two-tailed Wilcoxon rank-sum test) for FNI vs HC (purple) and SC vs HC (orange). *n.s.* stands for non-significant.

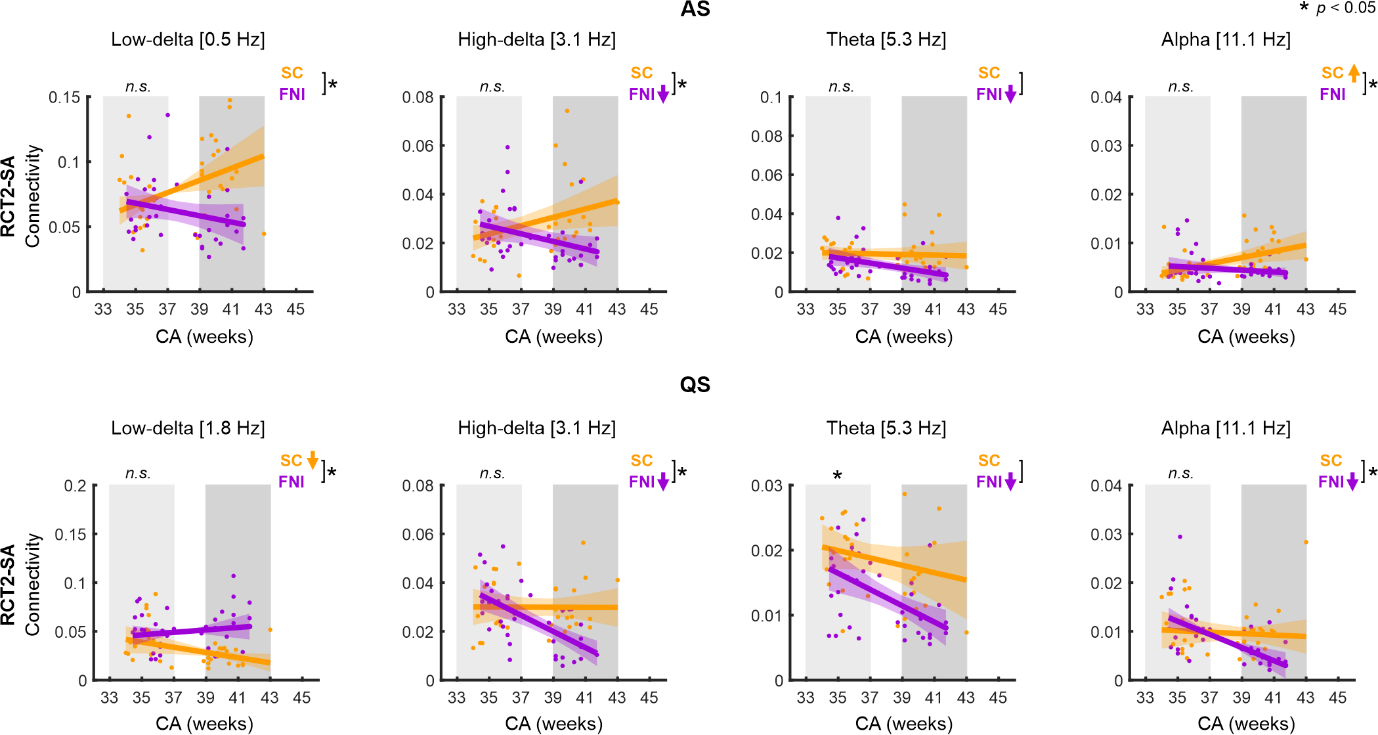

**Supplementary Fig. 4. Development of FNI-affected networks from 33 to 45 weeks CA in RCT2-SA.** Mean network strength is presented as a function of CA in the FNI (purple) and SC (orange) groups for the RCT2-SA cohort in AS (top) and QS (bottom) with confidence intervals shaded in the same color. Grey shaded areas depict the early preterm age (light grey) and term age (dark grey) windows. Group difference (two-tailed Wilcoxon rank-sum test) was computed between FNI and SC groups in the preterm age window. n.s. stands for non-significant results after Benjamini-Hochberg correction. Correlation of network strength with age (Spearman correlation) was computed for both groups; next to each plot, arrows indicate significant (p < 0.05) increase (upwards) or decrease (downwards) of PPC strength with CA in each group. Asterisks next to the square bracket indicate a significant (p < 0.05) difference in the correlation coefficients of the groups (two-tailed independent groups correlation of correlations, *cocor*).
